# T1DMicro: A Clinical Risk Calculator for Type 1 Diabetes Related Microvascular Complications

**DOI:** 10.1101/2021.09.01.21262996

**Authors:** Paul Minh Huy Tran, Eileen Kim, Lynn Kim Hoang Tran, bin Satter Khaled, Wenbo Zhi, Shan Bai, Diane Hopkins, Melissa Gardiner, Jennifer Bryant, Risa Bernard, John Morgan, Bruce Bode, John Chip Reed, Jin-Xiong She, Sharad Purohit

## Abstract

Development of complications in type-1 diabetes patients can be reduced by modifying risk factors. We used a cross-sectional cohort of 1646 patients diagnosed with type 1 diabetes (T1D) to develop a clinical risk score for diabetic peripheral neuropathy (DPN), autonomic neuropathy (AN), retinopathy (DR), and nephropathy (DN). Of these patients, 199 (12.1%) had DPN, 63 (3.8%) had AN, 244 (14.9%) had DR, and 88 (5.4%) had DN. We selected five variables to include in each of the four microvascular complications risk models: age, age of T1D diagnosis, duration of T1D, and average systolic blood pressure and HbA1C over the last three clinic visits. These variables were selected for their strong evidence of association with diabetic complications in the literature and because they are modifiable risk factors. We found the optimism-corrected R2 and Harrell’s C statistic were 0.39 and 0.87 for DPN, 0.24 and 0.86 for AN, 0.49 and 0.91 for DR, and 0.22 and 0.83 for DN respectively.

This tool (https://ptran25.shinyapps.io/Diabetic_Peripheral_Neuropathy_Risk) was built to help inform patients of their current risk of microvascular complications and to motivate patients to control their HbA1c and systolic blood pressure in order to reduce their risk of these complications.

## INTRODUCTION

The hyperglycemic state present in type 1 diabetes is associated with both micro- and macro-vascular complications [1; 2]. Microvascular damage leads to neuropathy, retinopathy, and nephropathy, which are each associated with clinical sequelae. Diabetic peripheral neuropathy (DPN) can lead to poor wound healing, diabetic ulcers, and eventually amputation [3]. Autonomic neuropathy (AN) can present with cardiac abnormalities, gastroparesis, or erectile dysfunction [4]. Diabetic retinopathy (DR) can lead to blindness [5]. Diabetic nephropathy (DN) can progress to end stage renal disease, requiring dialysis or renal transplantation [6].

Microvascular complications are major predictors of macrovascular complications, like myocardial infarctions and cerebrovascular accidents, which are the leading cause of death in the USA [7]. The most effective method to reduce morbidity and mortality in diabetic patients is minimizing the risk of macrovascular complications. This involves 1) identifying modifiable risk factors for developing microvascular complications, 2) motivating patients to reduce their personal risks, and 3) providing patients with tools to achieve risk reduction.

One of the most important modifiable risk factors is glycemic control. Multiple randomized control trials have demonstrated that tight glycemic control is associated with decreased risk of micro- and macro-vascular complications [8; 9]. Other factors have been associated with risk of microvascular complications in cross-sectional and longitudinal studies including sex [10], onset of type 1 diabetes during puberty [11], glycemic variability [12], quality of life during adolescence [13], and hypertension [14].

Once modifiable risk factors are identified, clinicians can educate patients on how to reduce these risk factors through lifestyle changes and medical management. Informing patients of their personal risks for developing diabetic complications helps patients set a realistic understanding of these risks and allows them to monitor how their lifestyle and/or medication changes have reduced their risks of diabetic complications [15]. Particularly for patients who already suffer from a lifelong, time-consuming, and usually expensive disease, risk scores can help prioritize their future-problem mitigation plan.

Risk scores have been reported in the past for complications of type 1 [16; 17; 18; 19] and type 2 diabetes [20]. Kazemi et al. published a support vector machine model using 13 clinical variables to predict DPN severity with an accuracy of 76% [17]. Lagani et al. used an accelerated failure model on the Diabetes Control and Complications Trial (DCCT) data to predict time to DPN onset using five variables—HbA1C, albumin, age, degree of retinopathy, and duration of post-pubescent diabetes—with a concordance index of 0.74 on a test data set [18]. They similarly used an accelerated failure model to predict time to retinopathy using five variables—HbA1c, marital status, degree of retinopathy, post pubescent diabetes duration, and body mass index—with concordance level of 0.72 on a test data set. Their random survival forest model for time to microalbuminuria based on six variables—HbA1c, marital status, urine albumin value, insulin regime, degree of retinopathy, post pubescent diabetes duration, and weight—had a concordance level of 0.82 on a test data set. DCCT modeled DPN risk using a generalized estimating equation with the variables mean HbA1c, age, height, duration of T1D, presence of DR, urinary albumin excretion rate, mean heart rate, and use of beta blocker. Braffett et al. also used a generalized estimating equation to model cardiovascular AN risk with the variables age, urinary albumin excretion rate, HbA1c, duration of T1D, mean pulse, beta blocker use, systolic blood pressure (SBP), presence of diabetic retinopathy, macular edema, estimated glomerular filtration rate (eGFR) less than 60, and cigarette smoking status [19]. A risk score was developed for blindness and limb amputation in individuals with type 1 or type 2 diabetes based on cox proportional hazards models [20].

These risk scores have not been implemented in clinics due to several factors. First, these scores use complex statistical approaches that are not easily accessible for patients and clinicians to use for risk calculations. Second, the scores for each complication use different clinical variables, making it more difficult for patients to collect all of the data necessary for computing clinical risk for each complication. Third, these scores do not show patients how changing their modifiable risk factors would change their risk of developing diabetic complications. Scores should be easily accessible and easy to use so that patients can use them to monitor their progress [15]. Learning this risk may motivate patients to progress on the Prochaska and DiClemente stages of change [21].

With the increasing incidence and survival of patients with T1D [22; 23; 24], these complications are becoming more important to study. We used a cross-sectional study, Phenome and Genome of Diabetes Autoimmunity (PAGODA), to develop a clinical risk score for DPN, AN, DR, and DN in patients with T1D.

## METHODS

### Study population

Individuals diagnosed with T1D who attended the Augusta University (AU) Medical Center and/or endocrinology clinics in Augusta and Atlanta areas of Georgia between 2002 and 2010 were recruited into the Phenome and Genome of Diabetes Autoimmunity (PAGODA) study [25; 26]. For consented patients, demographic and clinical variables, including age, sex, date of T1D diagnosis, medical diagnoses, blood pressure, laboratory measurements, and medications were extracted for the last three clinic visits (**Table 1**). Diagnoses of DPN, AN, and DR were extracted from patients’ electronic health records. DN was diagnosed by the physician/endocrinologist based on the last three microalbumin/creatinine ratio (MACR) values. We used MACR>30 for the diagnosis of DN. The vast majority of subjects were diagnosed with DPN based on a neurological history and exam by the treating endocrinologist. Many, but not all, had further evaluations and confirmation of DPN by a neurologist. DR was diagnosed by the yearly screening fundoscopic exam and patients with concerning findings were referred to a ophthalmologist for diagnosis and treatment. The research was carried out according to The Code of Ethics of the World Medical Association (Declaration of Helsinki, 1997). All study participants gave written informed consent. The study was reviewed and approved by the institutional review board at AU.

**Table 1.**
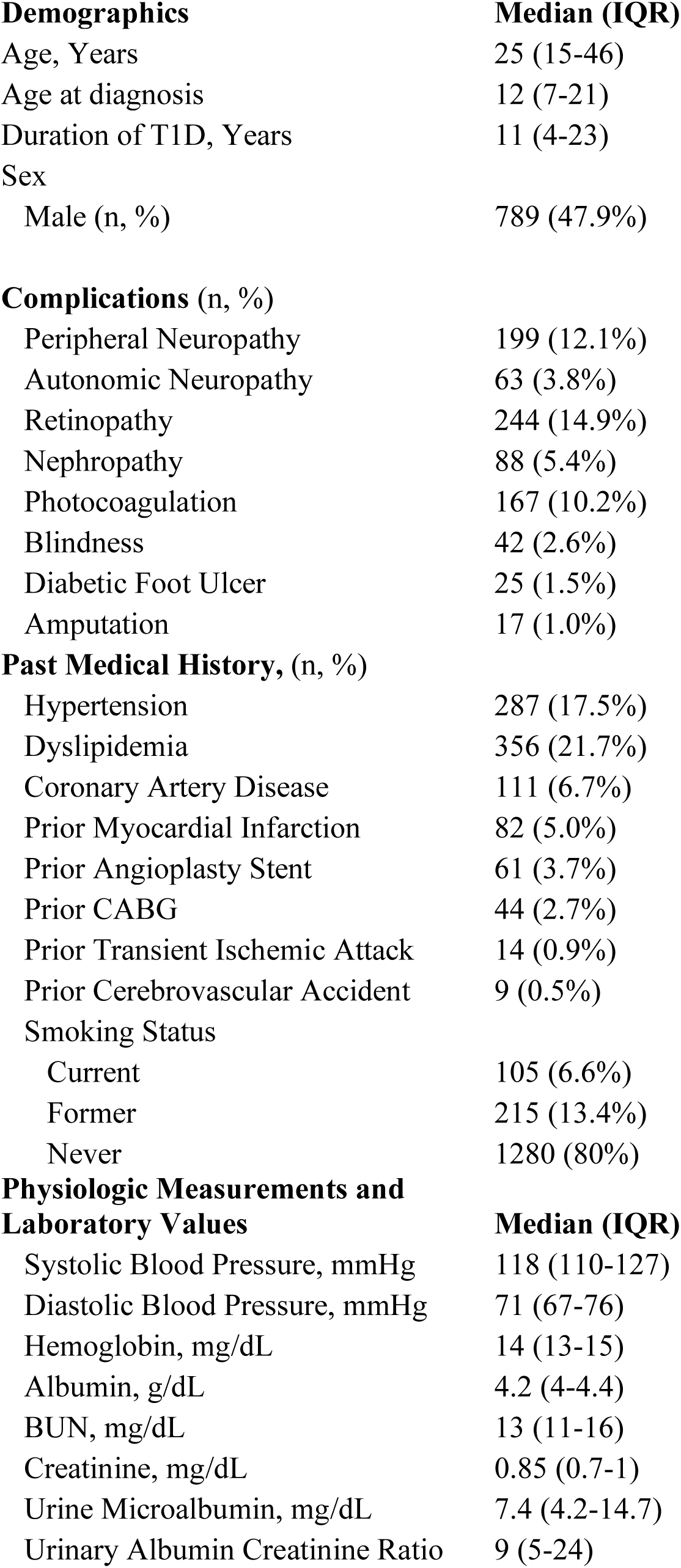
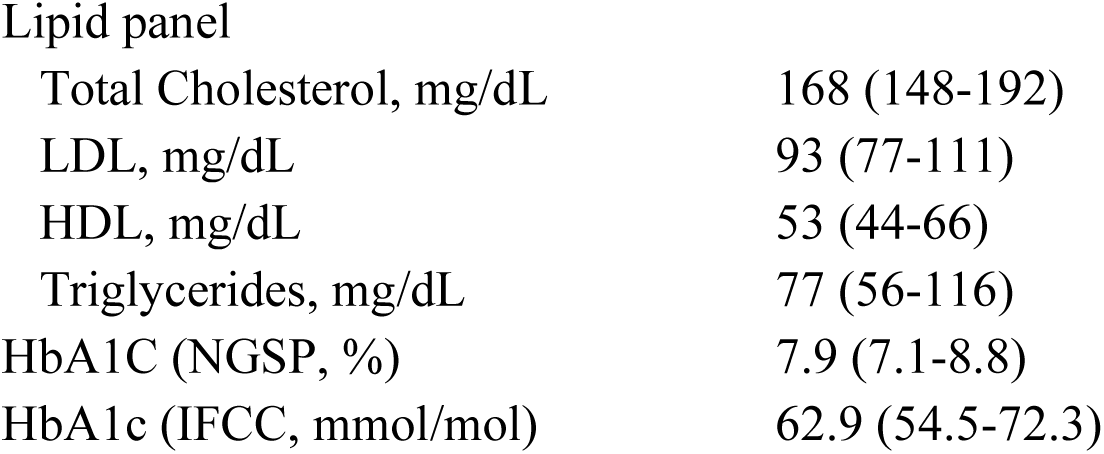
Demographics and Clinical Data of PAGODA subjects

| **Demographics** | **Median (IQR)** |
| --- | --- |
| Age, Years | 25 (15-46) |
| Age at diagnosis | 12 (7-21) |
| Duration of T1D, Years | 11 (4-23) |
| Sex |  |
| Male (n, %) | 789 (47.9%) |
| **Complications (n, %)** |  |
| Peripheral Neuropathy | 199 (12.1%) |
| Autonomic Neuropathy | 63 (3.8%) |
| Retinopathy | 244 (14.9%) |
| Nephropathy | 88 (5.4%) |
| Photocoagulation | 167 (10.2%) |
| Blindness | 42 (2.6%) |
| Diabetic Foot Ulcer | 25 (1.5%) |
| Amputation | 17 (1.0%) |
| **Past Medical History, (n, %)** |  |
| Hypertension | 287 (17.5%) |
| Dyslipidemia | 356 (21.7%) |
| Coronary Artery Disease | 111 (6.7%) |
| Prior Myocardial Infarction | 82 (5.0%) |
| Prior Angioplasty Stent | 61 (3.7%) |
| Prior CABG | 44 (2.7%) |
| Prior Transient Ischemic Attack | 14 (0.9%) |
| Prior Cerebrovascular Accident | 9 (0.5%) |
| Smoking Status |  |
| Current | 105 (6.6%) |
| Former | 215 (13.4%) |
| Never | 1280 (80%) |
| **Physiologic Measurements and Laboratory Values** | **Median (IQR)** |
| Systolic Blood Pressure, mmHg | 118 (110-127) |
| Diastolic Blood Pressure, mmHg | 71 (67-76) |
| Hemoglobin, mg/dL | 14 (13-15) |
| Albumin, g/dL | 4.2 (4-4.4) |
| BUN, mg/dL | 13 (11-16) |
| Creatinine, mg/dL | 0.85 (0.7-1) |
| Urine Microalbumin, mg/dL | 7.4 (4.2-14.7) |
| Urinary Albumin Creatinine Ratio | 9 (5-24) |

|  |  |  |
| --- | --- | --- |
| Lipid panel |  |  |
| Total Cholesterol, mg/dL |  | 168 (148-192) |
| LDL, mg/dL |  | 93 (77-111) |
| HDL, mg/dL |  | 53 (44-66) |
| Triglycerides, mg/dL |  | 77 (56-116) |
| HbA1C (NGSP, %) |  | 7.9 (7.1-8.8) |
| HbA1c (IFCC, mmol/mol) |  | 62.9 (54.5-72.3) |

### Statistical Analysis

The potential differences between T1D patients with and without each complication (DPN, AN, DR, and DN) were initially examined using univariate logistic regression. We selected five variables that showed consistent associations with diabetic complications across multiple studies and were modifiable through lifestyle and medication management: age, age at T1D diagnosis, duration of T1D, and average HbA1C and SBP over the last three clinic visits. These five variables were used to construct four multiple logistic regression models. Each model produced a clinical risk score for each diabetic complication in a microvascular naive T1D patient.

We determined the linearity of the relationship between each continuous variable and the microvascular complication using spiked histograms for visual analysis and analysis of variance of restricted cubic spline fits of the data for the statistical test of linearity. We used the spiked histograms to pre-specify the number of knots used for restricted cubic splines appropriate for each variable. The knots are placed at equal intervals across the distribution of the variables. These variables were all modeled with restricted cubic splines with 3, 5, and 4 knots, respectively. For the DR model, duration of T1D was also modeled with a restricted cubic spline with 3 knots. Calibration plots and validation were performed using the “calibrate” and “validate” functions, respectively, in the “rms” package [27] with 500 iterations of bootstrapping [28].

All p-values were two-tailed and a p<0.05 was considered statistically significant. All statistical analyses were performed using the R language and environment for statistical computing (R version 3.6.1; R Foundation for Statistical Computing; www.r-project.org). All data and code used to generate models, plots, and the website are available at https://github.com/pmtran5884/T1D_Complications.

## RESULTS

### Rates of diabetic complications

We consented a cross sectional cohort of 1646 patients diagnosed with T1D. Of the 1646 patients, 199 (12.1%) were diagnosed with diabetic peripheral neuropathy (DPN), 63 (3.8%) were diagnosed with autonomic neuropathy (AN), 244 (14.9%) were diagnosed with diabetic retinopathy (DR), and 88 (5.4%) were diagnosed with diabetic nephropathy (DN). Of these, 25 (1.5%) had diabetic foot ulcers and 17 (1%) had limb amputations. Of the patients with DR, 167 (10.2%) have had photocoagulation and 42 (2.6%) were blind. All patients were Caucasian. All clinical and demographic variables are listed in **Table 1**.

### Individual risk factors associated with diabetic complications

Univariate logistic regression analyses included several statistically significant results (Table 2). Not surprisingly, all four complications were associated with age, age at T1D diagnosis, and duration of T1D, as well as average SBP, DBP, hemoglobin, and blood urea nitrogen; dyslipidemia, and a macro-vascular condition (coronary artery disease, myocardial infarction, or cerebrovascular accident).

**Table 2.**
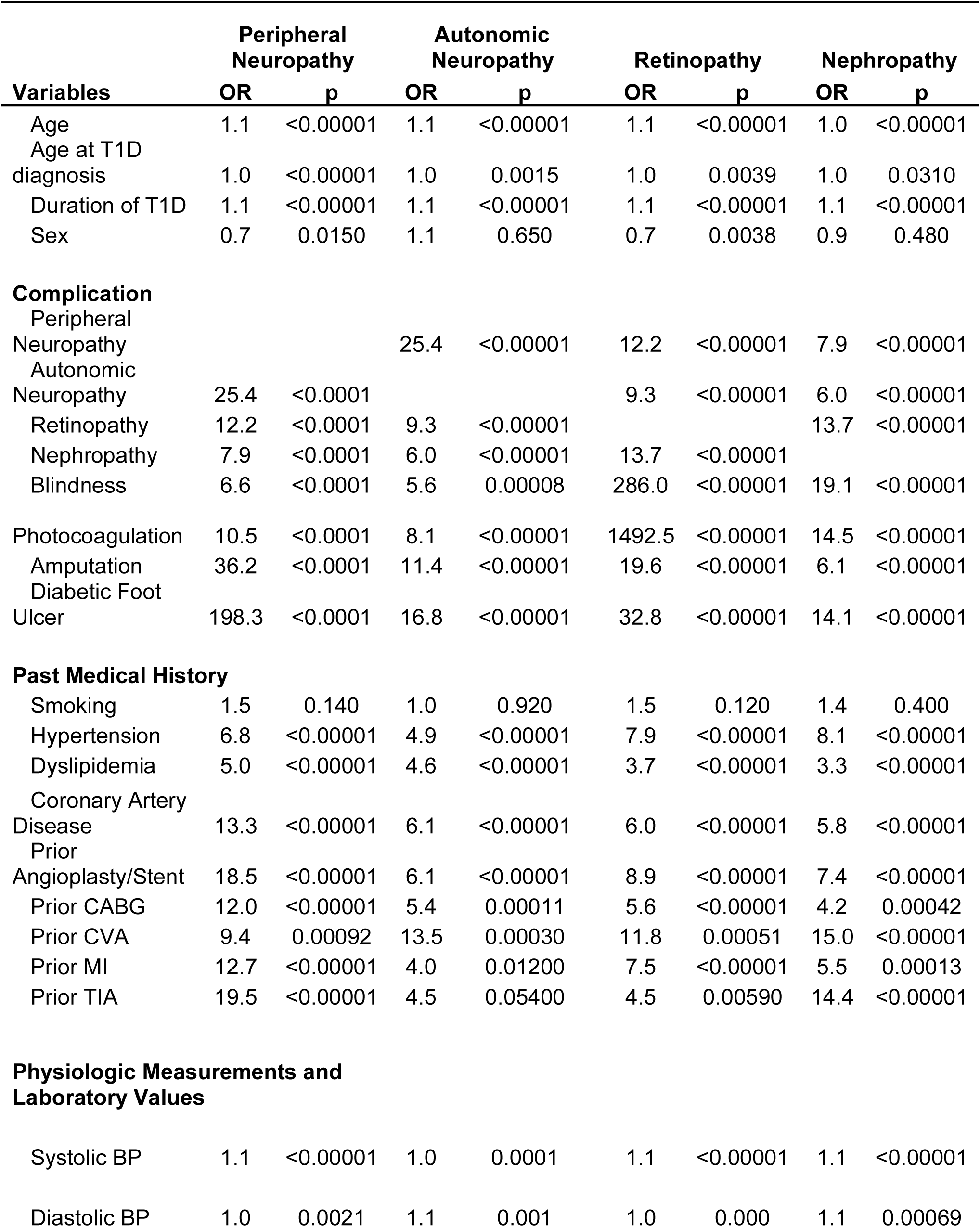
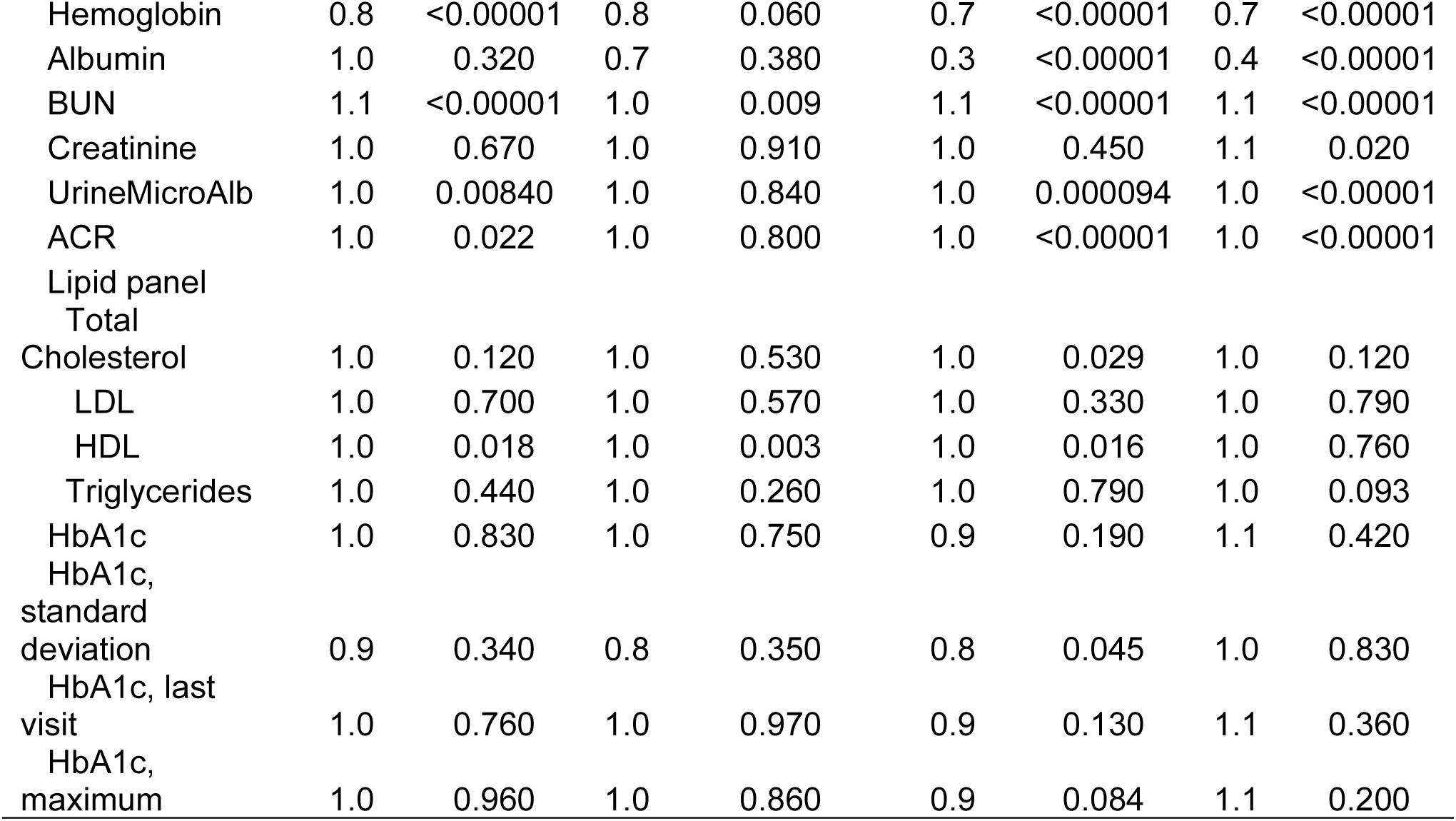
Univariate association of clinical variables with diabetic complications

Females were more likely to have DPN and DR than males consistent with previous studies [29]. Total cholesterol was associated with DR as well as high-density lipoprotein (HDL) but not low-density lipoprotein (LDL) nor triglycerides.

Interestingly, smoking status was not associated with any diabetic microvascular complications despite well-established associations between smoking and vascular disease. History of transient ischemic attack and microalbuminuria were associated with DPN, DR, and DN, but not AN; AN had the fewest significant associations with the patient variables in our analyses. Also interesting was that HbA1C was not statistically significantly associated with any of the four complications (average, maximum, or most recent) despite being the mainstay marker of diabetes severity.

### Multivariate predictive model of complications

We developed models to determine the risks of a microvascular complication naïve T1D patient for developing DPN, AN, DR, and DN based on five variables: patient age, duration of T1D, age at diagnosis of T1D, systolic blood pressure, and HbA1c. Spiked histograms demonstrated a non-linear relationship between current age, age at diagnosis of T1D, and average HbA1c over 3 clinic visits and the four diabetic complications.

In order to decrease the model complexity, we reasoned that variability accounted for by duration of T1D may be explained by current age and age of T1D onset. Thus, we compared models with and without this variable. The models without T1D duration were comparable to the models with this variable based on the likelihood ratio test for DPN (p=0.28) and AN (p=0.28), but performed worse for DR (p=3×10^×4^) and DN (p=0.04). We compared the calibration plots for each pair of models and found the mean squared error of the bias adjusted curves were similar for the DPN (2.3×10^×4^ with T1D duration, 2.2×10^×4^ without T1D duration), AN (4×10^×5^ and 6×10^×5^, respectively), and DN (8×10^×5^ and 2.4×10^×4^, respectively). We found the removal of the duration of T1D term gave an acceptable trade-off between model predictions and model interpretability for the DPN, AN, and DN models. The DR model retained the duration of type I diabetes term.

The models were validated using 500 iterations of bootstrapping with replacement. We found the optimism corrected R2 and Harrell’s C statistics were 0.39 and 0.87 for DPN, 0.24 and 0.86 for AN, 0.49 and 0.91 for DR, and 0.22 and 0.83 for DN. The calibration plot for each final model was generated from 500 iterations of bootstrapping with replacement and is presented in **Figure 1 (left)**. Calibration plots demonstrate all models are slightly overfit in the higher risk probability end of the models, with the AN and DN models slightly more overfit than the DPN and DR models.

**Figure 1.**
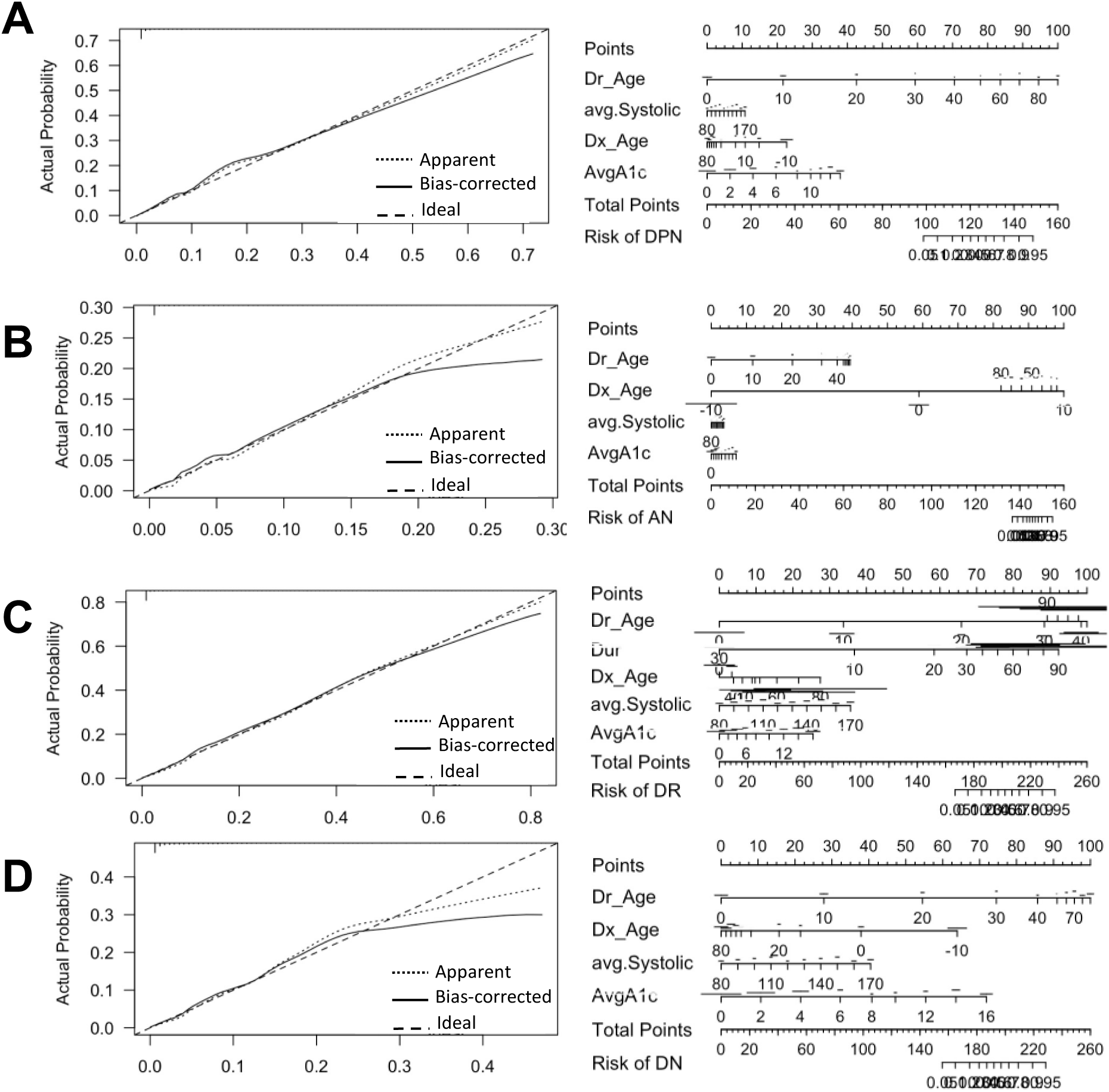
Calibration plot (left) and Nomogram (right) for DPN (A), AN (B), DR (C), and DN (D) in a multivariate logistic regression model. We applied the four microvascular complication models to the PAGODA dataset with 500 iterations of bootstrapping to generate the calibration plot showing the actual probability of complication in the y-axis and the predicted probability of complication based on the models in the x-axis. A bias correction was applied by calculating the difference in probability between the bootstrap iterations and the model prediction with the full dataset. The nomogram was generated again based on the beta coefficients from the four logistic microvascular complication models.

For both the DPN and AN models, the variables in order from most to least contributory were age, age at T1D diagnosis, average HbA1c, and average SBP. For the DR model, the variables in order from most contributory were age, average SBP, duration of T1D, age of T1D diagnosis, and average HbA1c. For the DN model, the variables in order from most contributory were age, age of T1D diagnosis, average SBP, and average HbA1c. Nomograms were generated as a visual aid for understanding variable contributions to the models (**Figure 1, right**).

### Web interface to predict individual risk of diabetic microvascular complications

To facilitate the use of our risk models by clinicians and patients, we created a web interface (https://ptran25.shinyapps.io/Diabetic_Peripheral_Neuropathy_Risk) for individuals to estimate their specific risks of diabetic microvascular complications. This interface was created to help inform patients of their personal risks of complications, motivate them to reduce their complication risk by reducing their SBP and HbA1C, and track their progress. In our models, changes in SBP and/or HbA1c levels were associated with noticeable changes in probability of having a microvascular complication (**Figure 2**). According to the International Society of Hypertension guidelines [30; 31], the target SBP for hypertensive patients is less than 140 mmHg. Our algorithm provides an additional risk estimate for patients whose SBP is greater than 140 mmHg had their SBP been 20 mmHg lower. According to the American Diabetes Association, International Society for Pediatric and Adolescent Diabetes, and Canadian Diabetes Association guidelines [32; 33; 34], the target HbA1C for diabetic patients is less than 7%. For patients whose HbA1C is greater than 7%, our algorithm also provides their risks had their HbA1C been 2% lower than entered. These additional risk scores for patients who have not met recommended SBP and HbA1C goals are intended to provide patients information about what their risks would be with improved blood pressure and blood glucose control.

**Figure 2.**
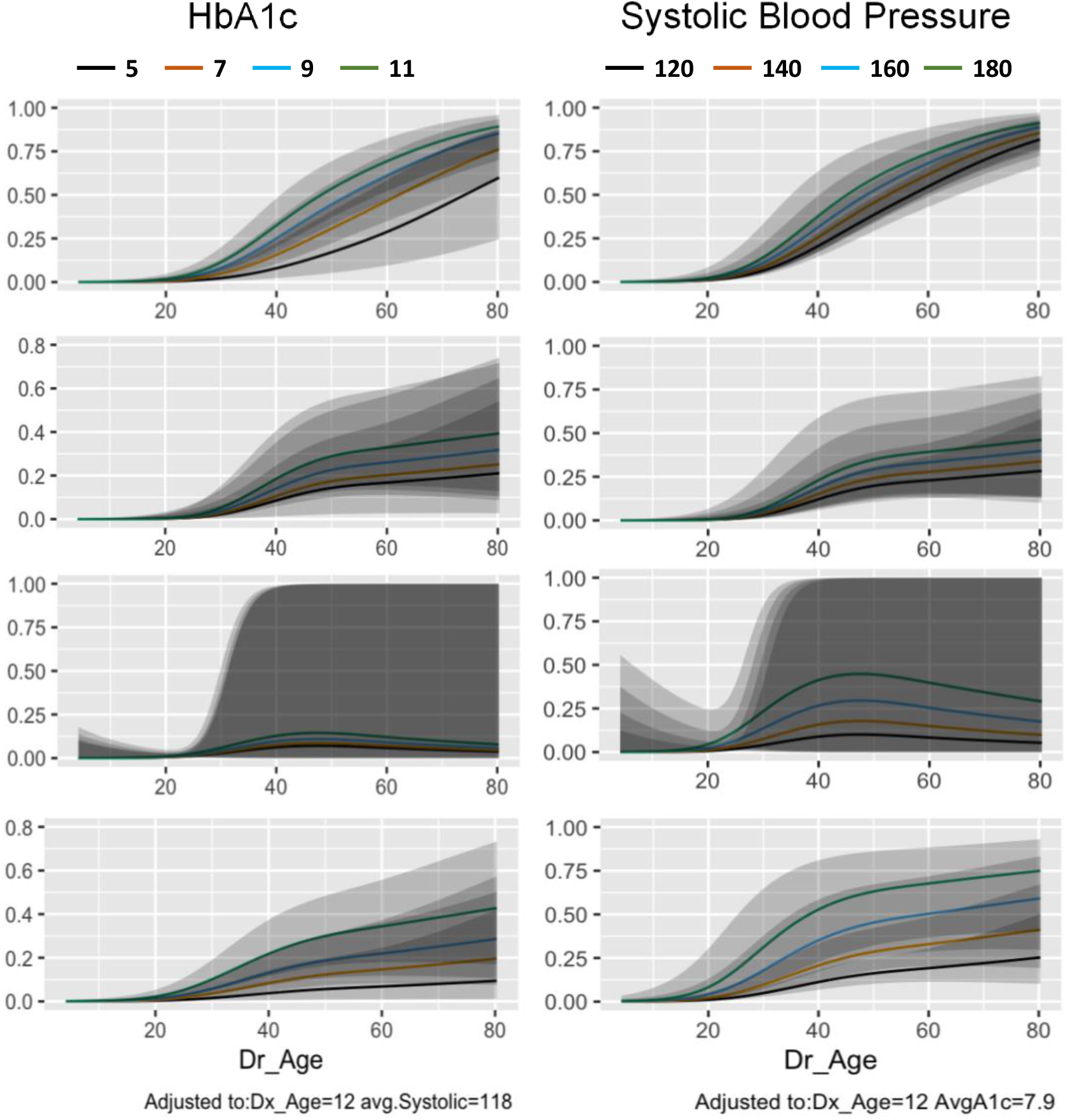
Predicted probability of DPN (A), AN (B), DR (C), and DN (D) with increasing HbA1c (left) and systolic blood pressure (right). We applied the four logistic microvascular complication models to simulated data by fixing the mean and standard deviation of the simulated data to that of the PAGODA population and varying the patient age at consent (Dr_Age) and our variables of interest, HbA1c and systolic blood pressure. We tested HbA1c at 5, 7, 9, and 11. We tested systolic blood pressure at 120, 140, 160, and 180. The gray zones show the 95% confidence interval of the predicted risk.

## DISCUSSION

While the DCCT trial and numerous other studies have shown the importance of HbA1C levels in the risk of diabetic complications [8; 9], our cross-sectional cohort had no association between any of the four diabetic complications and average HbA1c, maximum HbA1c, standard deviation of HbA1c, or most recent HbA1c on univariate analysis (**Table 2**). However, average HbA1c was a significant contributor in the multivariate logistic regression models (**Figure 1**), suggesting that HbA1c is important in the context of age and age at T1D diagnosis. Additionally, while the DCCT clinical trial compared glucose control through HbA1c in separate arms, our cross-sectional non-interventional cohort has lower HbA1c values on average and the HbA1c values were more closely distributed (**Table 1**). Additionally, we did not identify an association between smoking status and microvascular complications. It is possible that our patient population under-reported smoking or that the lack of pack years in our analysis led to the lack of statistical significance.

Most of the results from the univariate risk models agree with previous reports of risk factors for complications (**Table 2**). Similar to previous reports, we found a significant association between blood pressure, hypertension and the four diabetic complications [14]. We found that dyslipidemia was associated with diabetic complications [35]. We found that the association between microvascular complication and onset of T1D peaked around age 20. This is slightly older than the reported increased microvascular complication risk with T1D onset around puberty [11], but this may reflect a skew present in our cross-sectional cohort. Contrary to previous reports, we did not find any association between total cholesterol and diabetic complication risk [36].

We showed that the presence of one diabetic complication is strongly associated with having other diabetic complications (**Table 2**). This observation suggests that similar clinical variables may be used to predict multiple diabetic complications. Our models support this hypothesis since a logistic regression model including age, age at T1D diagnosis, average SBP, and average HbA1c performed well (**Figure 1**) with optimism-corrected R^2^ and Harrell’s C statistics 0.39 and 0.87 for DPN, 0.24 and 0.86 for AN, 0.49 and 0.91 for DR, and 0.22 and 0.83 for DN, respectively. This compares favorably to previously reported models [16; 17; 18; 19; 20] in terms of discrimination and calibration with the advantage of improved model simplicity and interpretability.

While other variables have been associated with T1D complications and applied in other risk models, we wanted to minimize the number of variables patients or clinicians need to identify to calculate complication risks. Namely, we did not include body mass index, triglycerides, or diastolic blood pressure [35]. Model simplicity might sacrifice the model’s discriminatory and calibration properties, but we have demonstrated that our models are comparable to previously reported models.

One of the limitations of this study is its cross-sectional nature. This does not allow for temporal analyses which can help to establish causation; for example, we are not able to discern if use of a statin or angiotensin converting enzyme inhibitor is associated with complications directly or via the diagnosis leading to use of the medication. The cross-sectional design also means we were unable to update our data with changes in diagnoses of complications, so our study likely underestimates the rates of diabetic microvascular complications. While we were able to address our models’ accuracy and calibration, validation is still required to establish generalizability. A third limitation due to the cross-sectional design is that the management of T1D including medications and screening for complications has invariably changed since patient recruitment ended in 2010, and we are not able to account for possible effects this might have had in predicting T1D complications.

Outside the above limitations owing to the cross-sectional design, the study population was entirely Caucasian. Although non-Hispanic Caucasians have the highest rate of T1D in the U.S., an estimated 23% of T1D is outside this demographic [37; 38]. Thus, it is possible that the results of our models are not generalizable to the entire T1D population. An additional limitation to generalizability is that we have not yet externally validated our models in an independent population, only via bootstrapping. Since the microvascular complication data were obtained from an electronic health record and not through regular screening, we would expect the complications to be underdiagnosed. Indeed, our calibration plots do all indicate the models predict a higher rate of diabetic complications compared to the training data.

We emphasize the important role of the modifiable factors HbA1c and SBP in the risk of developing microvascular complications (**Figure 2**) and informing patients of the potential reductions in risk associated with decreases in HbA1c and SBP. We hope this risk calculator becomes a useful tool for clinicians and patients and helps motivate patients to modify their risk factors for diabetic complications (https://ptran25.shinyapps.io/Diabetic_Peripheral_Neuropathy_Risk). The risk calculator is not intended to replace or support clinician diagnosis of microvascular complications. It is merely a tool to augment lifestyle counseling.

We characterized demographics, past medical history, blood pressure, laboratory values, and medications associated with four diabetic microvascular complications: DPN, AN, DR, and DN. We developed a clinical risk score for each microvascular complication using four clinical variables, age, age at T1D diagnosis, average SBP, and average HbA1C. The retinopathy model also included duration of T1D. We implemented this application as a web interface for clinicians and patients to easily calculate their risks of diabetic microvascular complications.

## Data Availability

All data are available upon reasonable request to authors. All code are available on the Github page.

https://github.com/pmtran5884/T1D_Complications

## AUTHOR CONTRIBUTIONS

PMHT, SP and JXS were involved with conception of the project. PMHT, LKHT and EK were responsible for data analysis; PMHT wrote the manuscript; WZ, BSK, SB, DH, MG, JB, RB, JM, BB, and JCR contributed to clinical samples. All authors contributed to writing and editing of the manuscript.

## ACKNOWLEDGEMENTS

We are very grateful to all patients and other volunteers who participated in this study.

